# Serial interval and generation interval for respectively the imported and local infectors estimated using reported contact-tracing data of COVID-19 in China

**DOI:** 10.1101/2020.04.15.20065946

**Authors:** Menghui Li, Kai Liu, Yukun Song, Ming Wang, Jinshan Wu

**Affiliations:** Beijing Institute of Science and Technology Information, Beijing, 100044, P. R. China; Beijing Science and Technology Information Strategy Decision-making Consultant Center, Beijing 100044, P. R. China; Faculty of Geographical Science, Beijing Normal University, Beijing, 100875, P. R. China; School of Systems Science, Beijing Normal University, Beijing, 100875, P. R. China

**Author notes:** Corresponding author Email address (Jinshan Wu).

**Keywords:** COVID-19, serial interval, generation interval, statistical analysis, contact tracing

## Abstract

**Backgrounds:** The emerging virus, COVID-19, has caused a massive outbreak worldwide. Based on the publicly available contact-tracing data, we identified 337 transmission chains from 10 provinces in China and estimated the serial interval (SI) and generation interval (GI) of COVID-19 in China.

**Methods:** Inspired by possibly different values of the time-varying reproduction number for the imported cases and the local cases in China, we divided all transmission events into three subsets: imported (the zeroth generation) infecting 1st-generation locals, 1st-generation locals infecting 2nd-generation locals, and others transmissions among 2+ generations. The corresponding SI (GI) is respec-tively denoted as 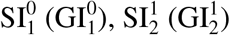, and 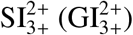. A Bayesian approach with doubly interval-censored likelihood is employed to fit the lognormal, gamma, and Weibull distribution function of the SI and GI using the identified 337 transmission chains.

**Findings:** It is found that the estimated 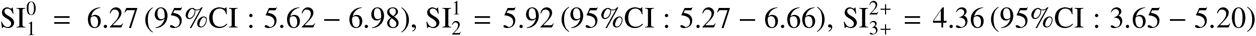, and 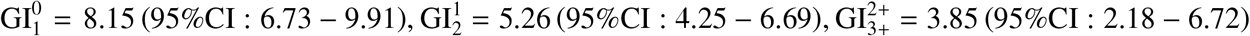, thus overall both SI and GI decrease when generation increases.

## 1. Introduction

As of April 11, 2020, COVID-19 has outbroken in 213 countries, areas or territories, and the World Health Organization (WHO) has reported over 1, 524, 161 confirmed cases and over 92, 941 confirmed deaths (WHO, 2020). It is a huge challenge to plan intervention strategies aimed at controlling outbreaks of COVID-19 in all countries, areas, or territories. Some key disease transmission parameters, including the basic reproduction number, the time-varying reproduction number, the generation interval (GI, time difference between being infected and infecting others), the serial interval (SI, the time difference between symptom onset of the infector and the infectee), and the incubation period (IP, the time difference between being infected and symptom onset), might offer insightful information of the epidemics and thus, might be helpful in devising interventions. In particular, the basic and time-varying reproduction numbers are good indicators of the speed of disease spreading and the effectiveness of interventions. Estimation of the basic and time-varying reproduction number often needs SI. In fact, for epidemics that are infectious during the incubation period, estimation of the reproduction numbers needs GI (Ganyani et al., 2020). It is possible that COVID-19 is infectious during incubation period (Qian et al., 2020, Wei et al., 2020). Therefore, in this work, we will perform a statistical analysis of both GI and SI.

As soon as investigation data were made available, several papers have quantified the GI, SI and IP of COVID-19 by employing statistical and mathematical modelling (Ganyani et al., 2020, Li et al., 2020, Zhao et al., 2020, You et al., 2020, Nishiura et al., 2020, Du et al., 2020, Tindale et al., 2020, Liu et al., 2020, Linton et al., 2020, Zhang et al., 2020, Backer et al., 2020, Bui et al., 2020, Moran, 2020, Wang & Teunis, 2020, Ping, 2020). Please see Table 1 for their estimated values and also the sample sizes. We find that the estimated values of SI from those previous studies are in a wide range: 3.95 − 7.5 days for SI. However, an accurate estimation of SI (and GI) is crucial in calculating the reproduction numbers accurately. Therefore, in this work, we firstly want to provide a more accurate estimation of SI (and GI) with possibly larger sample sizes. Secondly, if possible, we also want to shine some light on why there can be such larger differences in the estimated value of SI.

**Table 1:** Estimated values for serial interval, generation interval and incubation period in previous papers. *a* for Singapore, *b* for Tianjin, China. *:No amount of pairs given, this is just the number of cases in their dataset.

| Interval | Mean [95 CI%] | SD[95 CI%] | Samples(N) | Ref. |
| --- | --- | --- | --- | --- |
| SI | 3.95[-4.47-12.5] | 4.24[4.03-4.95] | 114<sup>b*</sup> | Ganyani et al. (2020) |
| SI | 3.96[3.53-4.39] | 4.75[4.46-5.07] | 486 | Du et al. (2020) |
| SI | 4.22[3.43-5.01] | - | 135<sup>b*</sup> | Tindale et al. (2020) |
| SI | 4.4[2.9-6.7] | 3.0[1.8-5.8] | 21 | Zhao et al. (2020) |
| SI | 4.41 | 3.17 | 71 | You et al. (2020) |
| SI | 4.56[2.69-6.42] | - | 93<sup>a*</sup> | Tindale et al. (2020) |
| SI | 4.7[3.7-6.0] | 2.9[1.9-4.9] | 28 | Nishiura et al. (2020) |
| SI | 4.8 | - | 112* | Wang & Teunis (2020) |
| SI | 5.1[1.3-11.6] | - | 35 | Zhang et al. (2020) |
| SI | 5.21[-3.35-13.94] | 4.32[4.06-5.58] | 91<sup>a*</sup> | Ganyani et al. (2020) |
| SI | 5.83 | 3.58 | 9 | Bui et al. (2020) |
| SI | 6.37 | 4.15 | 57 | Ping (2020) |
| SI | 6.6 | - | 12 | Moran (2020) |
| SI | 6.70[6.31-7.10] | 5.20[4.91-5.46] | 689 | Ma et al. (2020) |
| SI | 7.5[5.5-19] | 3.4 | 6 | Li et al. (2020) |
| GI | 5.2 [3.78-6.78] | 1.72[0.91-3.93] | 91<sup>a*</sup> | Ganyani et al. (2020) |
| GI | 3.95[3.01-4.91] | 1.51[0.74-2.97] | 114<sup>b*</sup> | Ganyani et al. (2020) |
| IP | 3.9 | - | 25 | Moran (2020) |
| IP | 4.8[2-11] | - | - | Liu et al. (2020) |
| IP | 5.0[4.2-6.0] | 3.0[2.1-4.5] | 52 | Linton et al. (2020) |
| IP | 5.2[1.8-12.4] | - | 49 | Zhang et al. (2020) |
| IP | 5.2[4.1-7.0] | - | 10 | Li et al. (2020) |
| IP | 6.4[5.6-7.7] | 2.3[1.7-3.7] | 88 | Backer et al. (2020) |
| IP | 7.1[6.13-8.25] | - | 93<sup>a</sup> | Tindale et al. (2020) |
| IP | 7.44[7.10-7.78] | 4.39[3.97-4.49] | 587 | Ma et al. (2020) |
| IP | 8.06[6.89-9.36] | - | 93 | Ping (2020) |
| IP | 9[7.92-10.2] | - | 135<sup>b</sup> | Tindale et al. (2020) |
| IP | 10.91 | - | 67 | You et al. (2020) |

Another motivation for this work comes from the extended framework of estimating the time-varying reproduction number of COVID-19 in China (Song et al., 2020). When working on determining the time-varying reproduction number of COVID-19 in China, we note that due to the different interventions for imported cases and local cases, their time-varying reproduction number should be different. Previously all analyses, as far as we know, have assumed that they are the same. See, for example, EpiEstim 2 (Thompson et al., 2019), which is a well-known R software on the estimation of time-varying reproduction number. For that, we need to distinguish the reported cases into the zeroth-generation imported cases *X*^0^, the first-generation locals infected by the imported cases *X*^1^, and so on, such as *X*^2^ and *X*^3+^. From the transmission chains among those cases, we then find SI and GI between various generations, such as 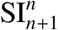 and 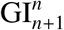, the SI and GI between the *n*th generation and the (*n* + 1)th generation.

There are three tasks in the above motivations, namely, obtaining more reliable estimates of GI and SI of COVID-19, finding out possible reasons for considerable differences in previous appraisals, and also providing GI and SI for various generations. To accomplish all the three tasks above, we extracted from online reports released by 10 provincial health commissions in China except Hubei. From that data, we identified 337 transmission chains and estimated transmission parameters. As we will see later, the SI and GI for various generations are indeed quite different.

## 2. Materials and Methods

We collected our data from publicly available official reports of case investigations published by provincial/municipal health commissions in China. The case investigations were performed by investigators in the corresponding centers for disease control and prevention in each province. The details of each confirmed case include the following necessary information: case ID, gender, age, date of symptom onset, date of diagnosis, history of traveling to or residing in Hubei or cities other than reporting city, date of arriving at the city where the case is reported. If identified via contact tracing performed by centers for disease control and prevention officers, the details also include contact case ID and date of exposure. The data includes 2300 confirmed cases that were compiled from online reports from 10 provinces in mainland China except Hubei between January 21, 2020, and February 29, 2020. Moreover, the cases are classified into different groups according to travel or residency history and chains of transmission of infection, if data on the case allows, as follows,

1. Imported cases (*X*^0^): Cases known to be infectors outside of Hubei but known to come out from Hubei recently,
2. Local first-generation cases (*X*^1^): Cases known to be infected by the imported cases,
3. Local second-generation cases(*X*^2^): Cases known to be infected by the local first-generation cases,
4. Local third-plus cases (*X*^3+^): Cases known to be infected by local second or higher generation cases.

Imported cases can be Hubei residents, or people traveled to Hubei very recently as long as they just came out from Hubei recently and became infectors in other provinces in China. We call the former Hubei residents and the latter Hubei travelers. Note that here Hubei residents are the ones who have been living in Hubei for a long time but recently came out from Hubei to another province in China. The date of symptom onset is defined as the appearing date of symptoms relevant to COVID-19. The exposure date, which is needed for estimating GI, is estimated to be the middle of data for the earliest and latest possible exposure time for local cases and also for Hubei travelers. For Hubei residents, their exposure date is hard to find due to our lack of data on Hubei cases. Therefore, whenever the exposure date is needed, we discard our data on Hubei residents. We processed the interval-censored data in units of days and discarded non-positive values, which means, in the case of SI, the infector show symptoms latter than the infectee. This assumption might well be the truth or due to some error in data collection, especially when the infector and the infectee are from the same household. We did find many cases with non-positive values are from the same household. We decide not to use those non-positive data since it is hard to tell who the infector is between the pairs in the same household. Finally, we obtained 337 COVID-19 transmission events, and we named this dataset as “All”. Then, we divided the “All” data into three subsets: Imported-first subsets 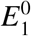, local first-second subsets 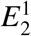, local second-third plus subsets 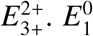 are composed of the events that imported cases *X*^0^ infect local first-generation cases *X*^1^, and others are defined accordingly. From these transmission chains, we get SI and GI for various generations. A Bayesian approach with doubly interval-censored likelihood (Reich et al., 2009) is then employed to obtain estimates of serial interval distribution, generation interval distribution, and incubation period distribution using the CmdStan (Nishiura et al., 2020) package in R.

## 3. Results

### 3.1 Serial interval

For all 337 samples, the observed SIs have a mean at *µ*_SI_ = 5.78 days and a standard deviation (SD) at *δ*_SI_ = 3.92 days. By using all these 337 samples, we estimated the mean at 5.80(95%CI : 5.38 − 6.24) days and SD at 3.95(95%CI : 3.57 − 4.40) days for gamma distribution. We also applied the estimation based on the lognormal distribution and the Weibull distribution. The fitted distributions are shown in Fig. 1 and the estimated parameters are reported in Table 2. We can see that, for most cases, the sample mean and sample SD agree quite well with the estimated values according to the gamma, lognormal, and Weibull distribution. From now on, in the main text, we only report sample values and fitted values from a gamma distribution.

**Table 2:** Estimated values of SI and GI. The widely applicable information criterion (WAIC) can be used to select model: The one with minimal WAIC value can be regarded as the best-fit model. Note that, for most cases, while the means of GI and SI are not the same, although still not that different either since they are often within their 95%CIs, their standard deviations are clearly different. Of course, for intervals upon arrival, GI and SI should be different in definition since SI^arrival^ − GI ^arrival^ ≈ IP > 0.

| Group |  | interval | Mean [95 CI%] | SD[95 CI%] | WAIC |
| --- | --- | --- | --- | --- | --- |
| All | lognormal | SI | 5.9(5.42, 6.45) | 4.90(4.18, 5.79) | 1778 |
|  |  | GI | 5.82(4.87, 6.99) | 3.24(2.25, 4.82) | 219 |
|  | gamma | SI | 5.80(5.38, 6.24) | 3.95(3.57, 4.40) | 1771 |
|  |  | GI | 5.96(5.0, 7.07) | 3.07(2.28, 4.21) | 218 |
|  | weibull | SI | 5.82(5.43, 6.24) | 3.86(3.52, 4.25) | 1776 |
|  |  | GI | 5.98(5.06, 6.98) | 2.83(2.23, 3.72) | 217 |
| Imported<br>-First | lognormal | SI | 6.38(5.62, 7.29) | 5.71(4.53, 7.27) | 925 |
|  |  | GI | 7.87(6.54, 9.45) | 2.73(1.76, 4.43) | 69 |
| | | $SI^{arrival}$ | 10.71(9.94, 11.55) | 5.73(4.93, 6.72) | 1165 |
| | | $GI^{arrival}$ | 2.68(2.14, 3.35) | 1.75 (1.14, 2.8) | 150 |
|  | gamma | SI | 6.27(5.62, 6.98) | 4.46(3.84, 5.19) | 917 |
|  |  | GI | 8.15(6.73, 9.91) | 2.85(1.84, 4.60) | 68 |
| | | $SI^{arrival}$ | 10.65(9.95, 11.40) | 5.08(4.51, 5.74) | 1154 |
| | | $GI^{arrival}$ | 2.81(2.21, 3.56) | 1.85(1.30, 2.71) | 157 |
|  | weibull | SI | 6.28(5.66, 6.96) | 4.25(3.72, 4.91) | 917 |
|  |  | GI | 8.14(6.83, 9.49) | 2.44(1.69, 3.66) | 68 |
| | | $SI^{arrival}$ | 10.67(9.97, 11.39) | 4.99(4.54, 5.51) | 1158 |
| | | $GI^{arrival}$ | 2.81(2.15, 3.60) | 2.0(1.49, 2.86) | 162 |
| Local<br>First-Second | lognormal | SI | 5.93(5.21, 6.78) | 4.33(3.39, 5.62) | 583 |
|  |  | GI | 5.02(4.14, 6.18) | 1.56(0.82, 2.9) | 61 |
|  | gamma | SI | 5.92(5.27, 6.66) | 3.7(3.11, 4.42) | 581 |
|  |  | GI | 5.26(4.25, 6.69) | 1.76(0.91, 3.28) | 63 |
|  | weibull | SI | 5.94(5.28, 6.65) | 3.61(3.12, 4.24) | 584 |
|  |  | GI | 5.16(4.26, 6.31) | 1.46(0.82, 2.56) | 60 |
| Local<br>Second-<br>Third+ | lognormal | SI | 4.24(3.54, 5.14) | 3.17(2.26, 4.4) | 262 |
|  |  | GI | 3.08(1.86, 4.72) | 2.41(1.06, 5.64) | 65 |
|  | gamma | SI | 4.36(3.65, 5.20) | 2.89(2.26, 3.77) | 266 |
|  |  | GI | 3.85(2.18, 6.72) | 3.27(1.34, 8.29) | 70 |
|  | weibull | SI | 4.38(3.65, 5.22) | 2.94(2.38, 3.77) | 270 |
|  |  | GI | 3.79(2.27, 6.36) | 2.74(1.26, 7.43) | 46 |

**Figure 1:**
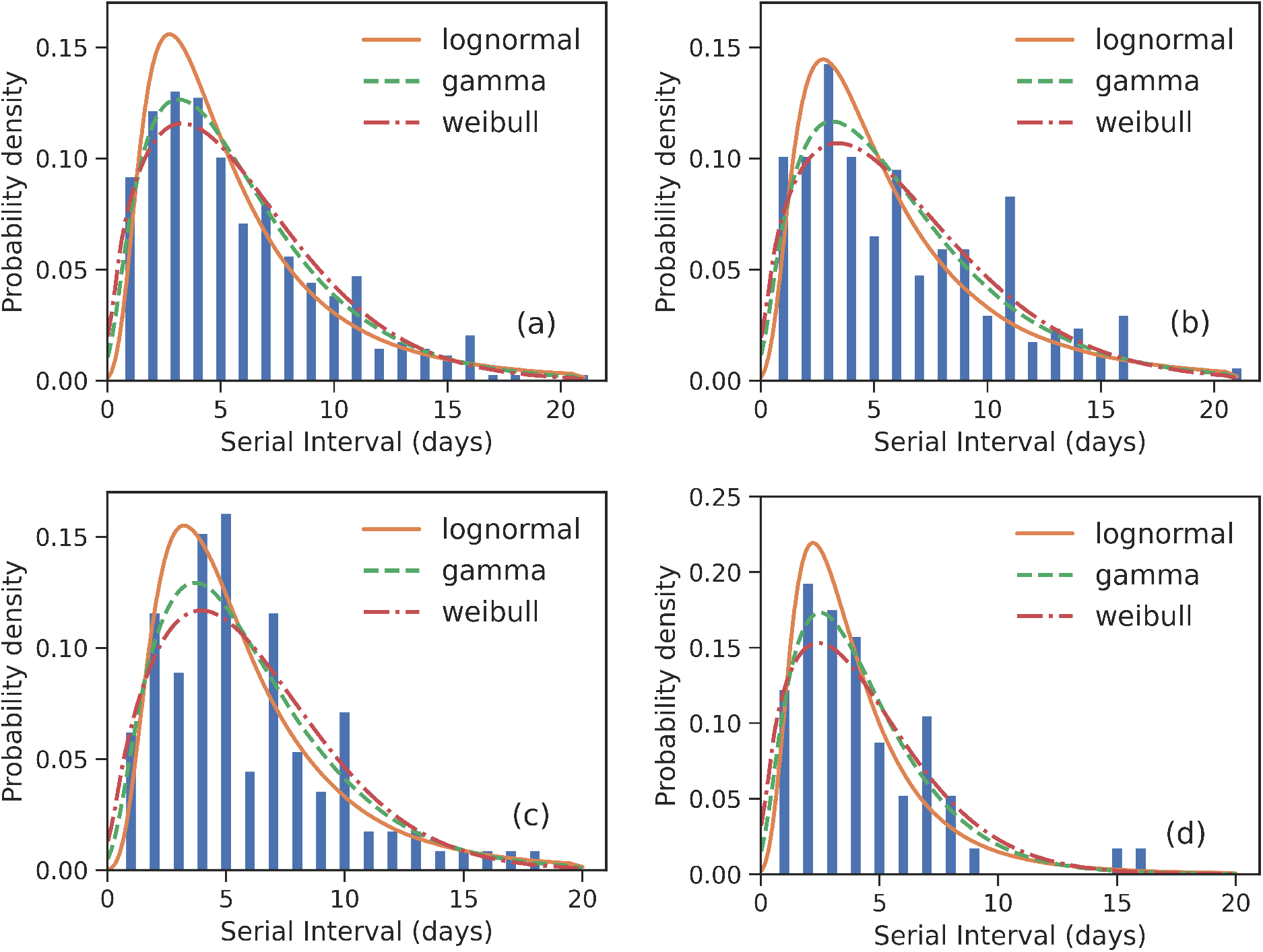
Fitted SI distribution for COVID-19 based on 337 reported transmission pairs in China between January 21, 2020 and February 29, 2020. Bars indicate the empirical distribution of SI samples and lines indicate the fitted lognormal, gamma and Weibull distributions respectively. (a) The “All” dataste (*N* = 337). (b) The imported-first subset 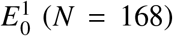. (c) The local first-second subset 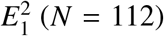. (d) The local second-third Plus subset 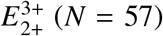. Values of the fitted parameters can be found in Tab. 2.

To further understand the wide range of the previously reported SIs, we estimated the distribution of SIs on three subsets. For the imported-first subset 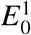 with 168 events, the observed SIs have a mean at *µ*_SI_ = 6.23 days and an SD at *δ*_SI_ = 4.24 days. We estimated the mean at 6.27(95%CI : 5.62− 6.98) and SD at 4.46(95%CI : 3.84 − 5.19) from the gamma distribution, as shown in Fig. 1 (b). Our estimated SI of the imported-first subset is slightly smaller than but close to the reported value of 7.5 in Li et al. (2020).

For the local first-second subset 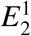 with 112 events, the observed SIs have a mean *µ*_SI_ = 5.88 days and a SD *δ*_SI_ = 3.66 days. We estimated the mean at 5.92(95%CI : 5.27− 6.66) days and SD at 3.70(95%CI : 3.11 −4.42) days from the gamma distribution as shown in Fig. 1 (c).

For the local second-third plus subset with 57 events, the observed SIs have a mean *µ*_SI_ = 4.28 days and a SD *δ*_SI_ = 3.05 days. we estimated the mean at 4.36(95%CI : 3.65 −5.20) days and SD at 2.89(95%CI : 2.26− 3.77) days for the gamma distribution as shown in Fig. 1 (d). The estimated SI is close to the lower bound 3.95 reported in Ganyani et al. (2020).

It is found that the estimated SI gradually decreases from 6.38 to 4.24 as generation increases. This discovery also explains to a certain degree why previous reported SIs in different papers are sometimes quite different. This result also reminds us to look into the reasons for such a trend in SI. Qian et al. (2020), Wei et al. (2020) pointed out that with more and more infective cases, it is more probable an earlier infection will happen if there are pre-symptomatic transmissions. The earlier infections will likely make SI smaller. Thus, the gradually decreasing SI leads us to examine whether or not there are pre-symptomatic transmissions.

### 3.2 Pre-symptomatic transmissions

To check if there are pre-symptomatic transmissions, we compared with the earliest exposure time of infectee and the onset time of infector. It is found that 95 of the 337 (28%) reports indicate that infectees may be infected before symptoms of infectors appear. Moreover, pre-symptomatic transmissions have occurred 36 of the 168 (21.4%) events in the imported-first subset, 34 of 112 (30.4%) events in the local first-second subset, 25 of 57 (43.9%) events in the local second-third plus subset. The ratio of pre-symptomatic transmission increases as generation increases.

### 3.3 Generation interval

GI distribution is needed for the inference of the reproduction number (Wallinga & Lipsitch, 2006). Often people use SI as a proxy of GI as the time of infection is not often reported in case files. In principle, SI and GI should have equal expected values since the IP time for the infector and infectee should cancel out. Consequently, GI is less studied than SI. However, firstly GI and SI still might have different standard deviations even if they have the same mean. As we will see later, it turns out that for COVID-19, even the means of GI and SI are slightly different, and their standard deviations are clearly different. Secondly and more importantly, for epidemics with pre-symptomatic transmissions, one needs GI instead of SI since, even before the onset of symptoms, transmissions can occur already. It has recently shown that estimates of the reproduction number are biased if ignoring the difference between SI and GI (Leonhard et al., 2019). Surprisingly, only very few papers have studied GI of COVID-19 (Ganyani et al., 2020). In this work, we would like to add one more study of GI of COVID-19.

To get a GI value, we need exposure times of both the infector and the infectee in a transmission chain. However, exposure time is not available for many cases. Especially for Hubei residents in the imported cases, due to our lack of contacttracing data in Hubei, it is impossible to know their exact exposure date accurately. Therefore, to estimate GIs, we only consider imported cases with travel history (i.e., to exclude the Hubei residents from the imported cases) and use the middle of their trips as their date of exposure since people can often remember much better the dates of their trips. After that, we only obtained 43 events for estimating GI from 337 transmission chains.

For the whole dataset with 43 events, the observed GIs have a mean of *µ*_GI_ = 6.40 days and an SD of *δ*_GI_ = 3.29 days. We estimated the mean at 5.96(95%CI : 5.0 − 7.07) days and SD at 3.07(95%CI : 2.28 − 4.21) days for the gamma distri-bution. The fitted distributions from the lognormal and Weibull distributions are shown in Fig. 2 and the estimated parameters are reported in Tab. 2.

**Figure 2:**
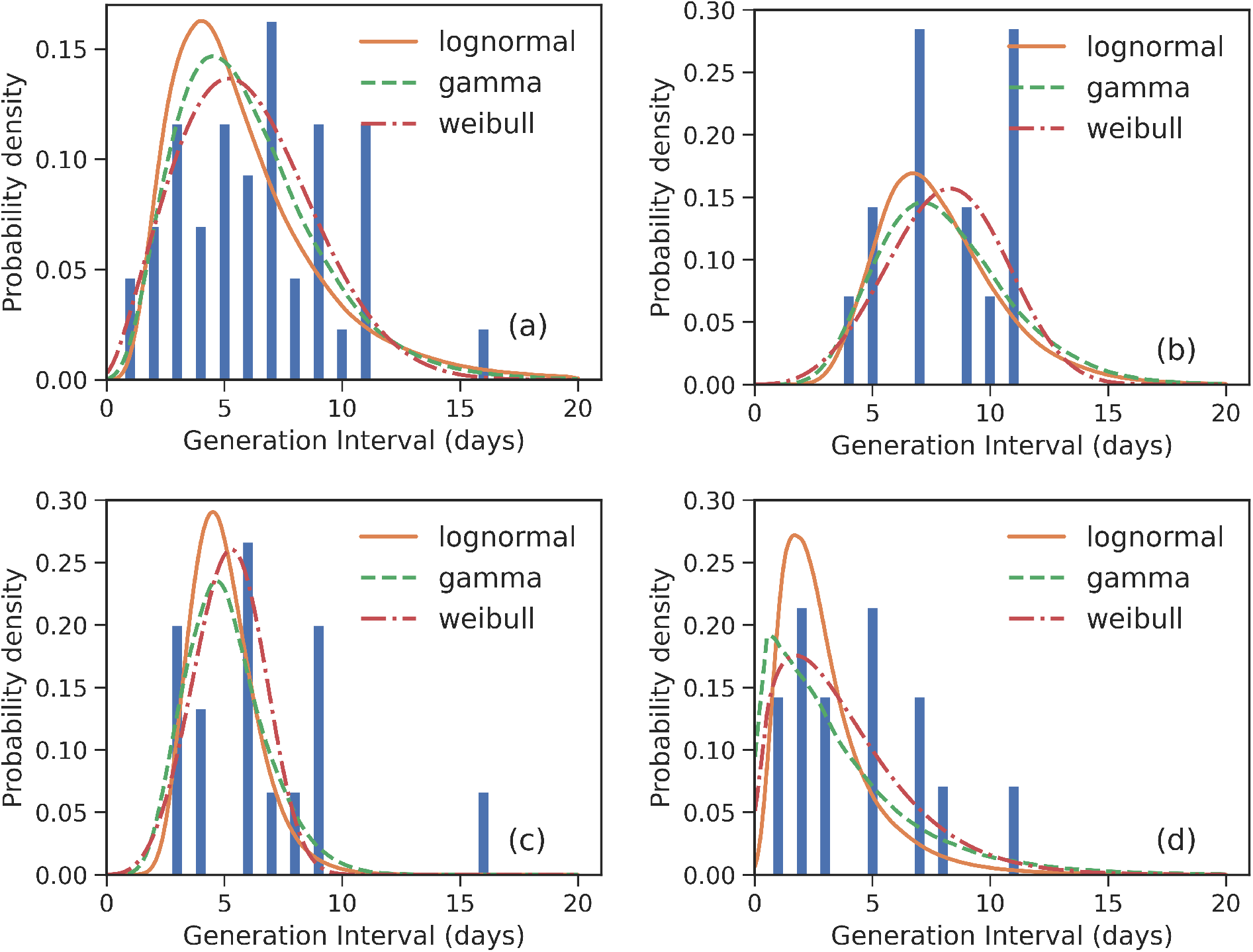
Fitted GI distributions for COVID-19 based on 43 reported transmission pairs. Bars indicate the empirical distribution of GI samples and lines indicate the fitted lognormal, gamma and Weibull distributions respectively. (a) The “All” dataste (*N* = 43). (b) The imported-first subset 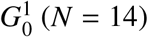. (c) The local first-second subset 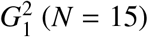. (d) The local second-third plus subset 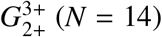. Values of fitted parameters can be found in Tab. 2.

For the imported-first subset with 14 events, the observed GIs have a mean of *µ*_GI_ = 8.14 days and an SD of *δ*_GI_ = 2.48 days. We estimated the mean at 8.15(95%CI : 6.73− 9.91) days and SD at 2.85(95%CI : 1.84 − 4.60) days for the gamma distribution.

For the local first-second subset with 15 events, the observed GIs have a mean *µ*_GI_ = 6.60 days and an SD of *δ*_GI_ = 3.40 days. We estimated the mean at 5.26(95%CI : 4.25 − 6.69) days and SD at 1.76(95%CI : 0.91 − 3.28) days for the gamma distribution.

For the local second-third plus subset with 14 events, the observed GIs have a mean *µ*_GI_ = 4.43 days and an SD of *δ*_GI_ = 2.98 days. We estimated the mean at 3.85(95%CI : 2.18 − 6.72) days and SD at 3.27(95%CI : 1.34 − 8.29) days for the gamma distribution.

The estimated mean values of GI and SI seem to be slightly different, although their confidence intervals overlap marginally. Their standard deviations are clearly different. Moreover, as the generation increases, the means of GIs decrease from 8.15 to 3.85. This is consistent with the decreasing SI, as reported in Sec. 3.1. Of course, such a difference between GI and SI may be caused by small sample size in our GI data, or they might be indeed different. This difference calls for further analysis, which in turn calls for more information to be provided in the reported case files. We would like to point out that such differences between GI and SI at least make it not suitable to replace the distribution of GI with the distribution of SI in estimating reproduction numbers, as noted already by Leonhard et al. (2019).

### 3.4 Incubation period

Depending on different sample datasets, the estimated incubation period (IP) in previous studies have an even wider ranged 3.9 − 10.91 days (Liu et al., 2020, Linton et al., 2020, Zhang et al., 2020, Li et al., 2020, Backer et al., 2020, Tindale et al., 2020, You et al., 2020, Moran, 2020, Ma et al., 2020, Ping, 2020). Please see Tab. 1 for their estimated values and sample sizes. Such a large discrepancy makes it difficult to plan for public health interventions.

To estimate IP, we need the date of exposure and the date of symptom onset for each case. We identify 545 cases satisfying this condition from our data. From all of our 545 samples, we observed *µ*_IP_ = 8.47 days and *δ*_IP_ = 5.0 days. We estimated the mean at 8.36(95%CI : 7.94 − 8.80) days and SD at 4.89(95%CI : 4.51 − 5.31) days for the gamma distribution. The fitted distributions from the lognormal and Weibull distributions are plot in Fig. 3 and the estimated parameters are reported in Tab. 3.

**Table 3:** Estimated IP values for various distributions and for various generations.

| Group |  | Mean [95 CI%] | SD[95 CI%] | WAIC |
| --- | --- | --- | --- | --- |
| All | lognormal | 8.43(7.95, 8.94) | 5.64(5.03, 6.35) | 3195 |
|  | gamma | 8.36(7.94, 8.80) | 4.89(4.51, 5.31) | 3189 |
|  | weibull | 8.38(7.96, 8.82) | 4.83(4.51, 5.19) | 3202 |
| Imported | lognormal | 7.4(6.87, 7.98) | 4.71(4.06, 5.50) | 1786 |
|  | gamma | 7.39(6.90, 7.91) | 4.2(3.76, 4.70) | 1787 |
|  | weibull | 7.41(6.92, 7.93 ) | 4.15(3.78, 4.58) | 1794 |
| Local<br>first<br>generation | lognormal | 8.97(8.07, 10.0) | 5.02(4.06, 6.32) | 605 |
|  | gamma | 8.94(8.11, 9.85) | 4.42(3.75, 5.26) | 598 |
|  | weibull | 8.94(8.15, 9.77) | 4.10(3.58, 4.76) | 595 |
| Local<br>second-plus<br>generation | lognormal | 10.19(8.95, 11.66) | 7.66(6.02, 9.90) | 764 |
|  | gamma | 10.15(9.03, 11.38) | 6.38(5.39, 7.61) | 757 |
|  | weibull | 10.19(9.11, 11.36) | 6.08(5.26, 7.13) | 756 |
| $IP^{arrival}$ | lognormal | 6.40 (6.03, 6.80) | 5.79(5.19, 6.48) | 4470 |
|  | gamma | 6.20 (5.90, 6.51) | 4.43(4.14, 4.75) | 4436 |
|  | weibull | 6.21(5.92, 6.51) | 4.30(4.05, 4.58) | 4443 |

**Figure 3:**
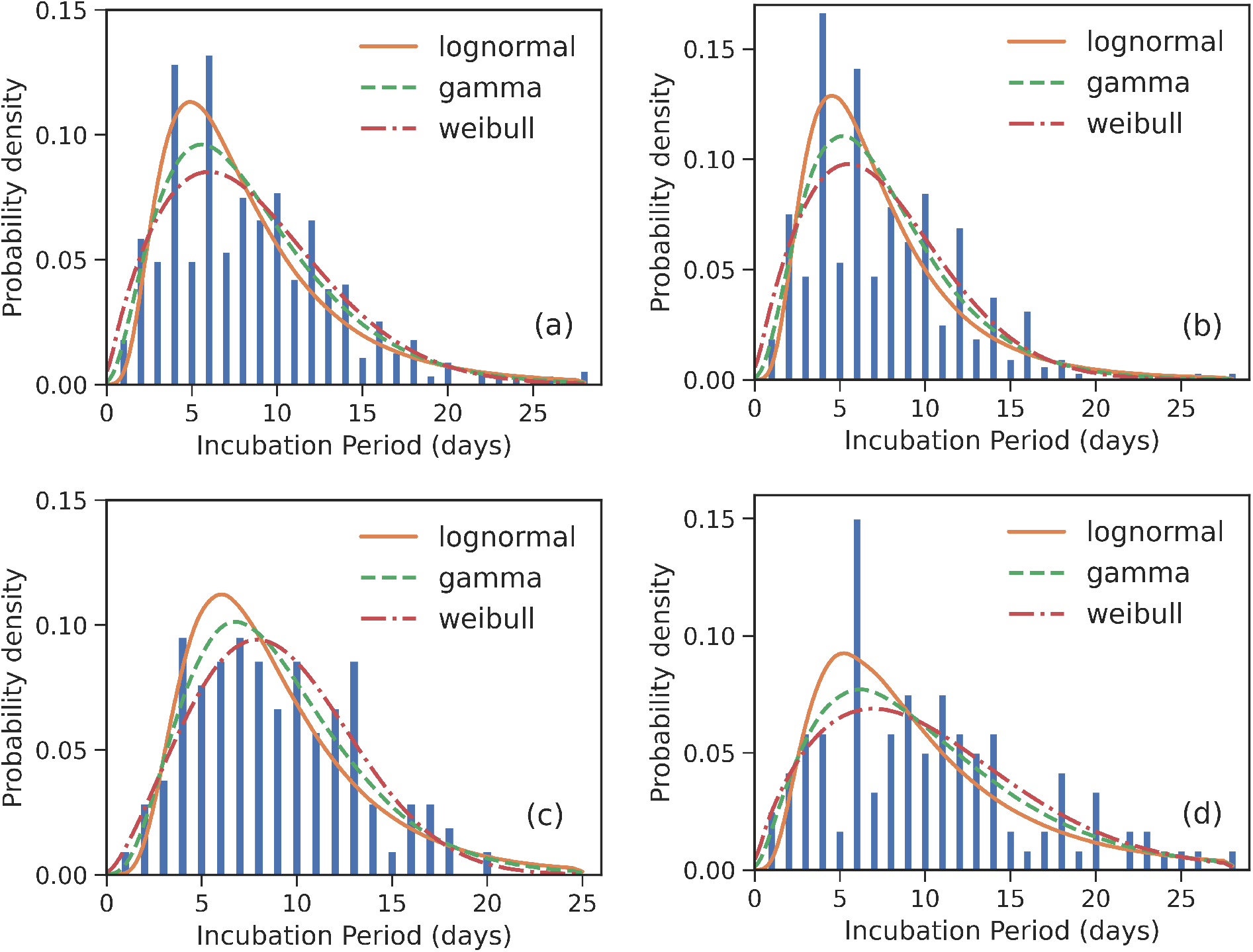
Fitted IP for COVID-19 based on 545 cases. Bars indicate the empirical distributions of IP samples and lines indicate the fitted lognormal, gamma and Weibull distributions respectively. (a) The “All” dataset (*N* = 545). (b) The imported cases subset (*N* = 320). (c) The local first-generation cases subset (*N* = 105). (d) The local second-plus cases subset (*N* = 120). Values of fitted parameters can be found in Tab. 3.

Again, we divide the dataset into three subsets, the imported cases with travel history (*X*^0,*T*^) (i.e., to exclude the Hubei residents from the imported cases), the local first-generation cases (*X*^1^), and the local second-plus generation cases (*X*^2+^). For the imported subset with 320 cases, the observed IPs have a mean of *µ*_IP_ = 7.69 days and an SD of *δ*_IP_ = 4.62 days. We estimated the mean at 7.39(95%CI : 6.90 − 7.91) days and SD at 4.2(95%CI : 3.76 − 4.70) days for the gamma distribution. We take the exposure date of the imported cases with travel history to be the middle of their trips since one can often remember dates of traveling accurately. Moreover, for most imported cases, their traveling times are often quite short.

For the local first-generation (*X*^1^) subset with 105 cases, the observed IPs have a mean *µ*_IP_ = 8.83 days and a SD of *δ*_IP_ = 4.20 days. We estimated the mean at 8.94(95%CI : 8.11 − 9.85) days and SD at 4.42(95%CI : 3.75 − 5.26) days for the gamma distribution.

For the local second-plus generation (*X*^2+^) subset with 120 cases, the observed IPs have a mean *µ*_IP_ = 10.22 days and a SD of *δ*_IP_ = 6.07 days. We estimated the mean at 10.15(95%CI : 9.03 − 11.38) days and SD at 6.38(95%CI : 5.39 − 7.61) days for the gamma distribution.

### 3.5. Intervals upon arrival for imported cases

Sometimes, for imported cases, in particular, knowing after their arrival typically how long they will show symptoms, infect locals, and also when the local infectees, who are infected by the imported cased, will show symptoms, can also be informative for decision-makers of intervention strategies. Therefore, in this work, we also show our results on these statistics.

The serial interval upon arrival (*S I*^arrival^) is defined as the interval between the date that an imported case arrives at the reporting city and the date that the infectee, infected by the imported case, shows symptoms. In our dataset, 194 transmission events meet this condition. The observed *S I*^arrival^ have a mean of *µ* _SI_^arrival^ = 10.63 days and a SD of *δ* _SI_^arrival^ = 4.98 days. We estimated the mean at 10.65(95%CI : 9.95− 11.4) days and SD at 5.08(95%CI : 4.51 − 5.74) days for gamma distribution, as shown in Fig. 4 (a). We also plot the fitted distributions from the lognormal and Weibull distributions in Fig. 4. The estimated *S I*^arrival^ is reported in Tab. 2.

**Figure 4:**
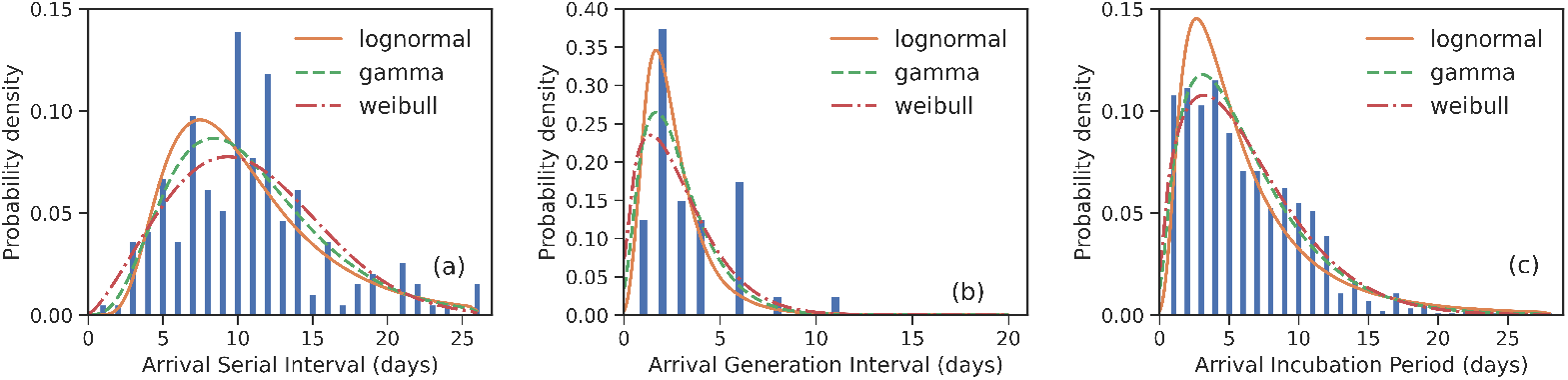
Fitted distributions of the various intervals upon arrival for imported cases for COVID-19. Bars indicate the empirical distributions and lines indicate the fitted lognormal, gamma and Weibull distributions respectively. (a) SI upon arrival (*S I*^arrival^) (*N* = 194). (b) GI upon arrival (*GI*^arrival^) (*N* = 40). (c) IP upon arrival (*IP*^arrival^) (*N* = 815). Values of fitted parameters are reported in both Tab. 2 and Tab. 3.

The generation interval upon arrival (*GI*^arrival^) is defined as the interval between the date that an imported case arrives at reporting city and the date that he/she infects others. In our dataset, 40 transmission events meet this condition. The observed *GI*^arrival^ have a mean of *µ*_GI_^arrival^ = 3.35 days and an SD of *δ*_GI_^arrival^= 2.19 days. We estimated the mean at 2.81(95%CI : 2.21 − 3.56) days and SD at 1.85(95%CI : 1.3 − 2.71) days for the gamma distribution as shown in Fig. 4 (b). The estimated *GI*^arrival^ is reported in Tab. 2. In definition, SI upon arrival is more or less the summation of GI upon arrival and IP, which is SI^arrival^ − GI^arrival^ ≈ IP > 0, unlike the relation between the usual SI and GI, SI ≈ GI.

The incubation period upon arrival (*IP*^arrival^) is defined as the interval between the date that an imported case arrives at the reporting city and the date that the imported case shows symptoms. In our dataset, 815 cases meet the above condition. The observed *IP*^arrival^ have a mean of *µ*_IP_^arrival^ = 6.19 days and an SD of *δ*_IP_^arrival^ = 4.37 days. We estimated the mean at 6.20(95%CI : 5.90 − 6.51) days and SD at 4.43(95%CI : 4.14 −4.75) days for the gamma distribution as shown in Fig. 4 (c). The estimated *IP*^arrival^ is reported in Tab. 3. It is found that *IP*^arrival^ is larger than *GI*^arrival^. This indicates again that pre-symptomatic transmissions do occur.

## 4. Conclusion and discussion

In this paper, we firstly estimated serial intervals (SI) based on 337 transmission events, which are divided into three subsets, including imported-first subset 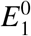, local first-second subset 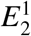 and local second-third plus subset 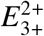. It is found that the estimated SI decreases with the number of generations and they are respectively 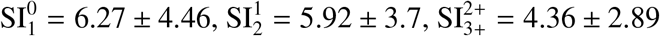. We also found that pre-symptomatic transmissions have likely occurred in 95 events out of 337 events (28%).

Then, we estimated the generation interval (GI) in the three subsets. It is also found that the estimated GI decreases as the generation increases, and they are respectively 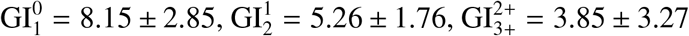. We would like to point out that there are small differences between the means of GI and the corresponding SI, and clear differences between their standard deviations. This, together with the existence of pre-symptomatic transmissions, makes it necessary to use GI in estimating reproduction numbers rather than SI.

Next, we estimated the incubation period (IP) of different groups of cases. It is found that the estimated IPs have a mean at IP^0,*T*^ = 7.39 ± 4.2 days for 320 imported cases with travel history, a mean at IP^1^ = 8.94 ± 4.42 for 105 local first-generation cases, and a mean at IP^2+^ = 10.15 ± 6.38 days for 120 local second plus-generation cases.

Moreover, we estimated the SI, GI, and IP upon arrival of the imported cases at the reporting city. It is found that the imported cases will show symptoms after IP^arrival^ = 6.19 ± 4.4 days arrival in reporting cities and will infect others after GI^arrival^ = 3.29 ± 2.65 days. The difference between these two intervals also indicates that pre-symptomatic transmission is likely to occur. Finally, it is found that the local first generating cases (infected by imported cases) will show symptoms after SI^arrival^ = 10.02 ± 4.91 days imported cases arrived at the reporting cities.

Providing statistics for various generation cases so that in further studies better models can be established, for example, by making use of different values of transmission parameters for different generations, is the main contribution of this work. Our results also explain to a certain degree that why in previous studies values of those estimated parameters span a wide range. For the imported cases, in particular, we reported SI, GI, and IP upon their arrivals. This study can be meaningful for both planning intervention and modeling epidemics. Furthermore, one should note that for epidemics with pre-symptomatic transmissions, in estimating the basic and the time-varying reproduction number, GI should be used instead of SI.

There are several limitations to this study. Our data is restricted to online reports from only 10 provinces in China. The content of epidemiological investigation reports from different provinces varies a lot. Many case reports do not have exposure date and infector ID, which are quite crucial in epidemics modeling. Thus, while admitting this limitation, here we also call for designing/utilizing a standard format of the case reports, countrywide, or even worldwide. Our sample size, especially on generation interval, is still very small. Thus, our results GI are not as reliable as the ones on SI.

## Data Availability

Data used for this study are all from publicly available sources. The data files are available upon request to the authors.

## 5. Acknowledgment

The authors would like to thank the data processing team, including Jiatong Zhu, Jianxin Zhang, Kaiwen Li, Yuting Zhang, Ningning Qiao, for their hard work of extracting the contact-tracing data from online reports. We should also note that it is the center of disease control and prevention of China at all levels, including national, provincial, county-level, and their great investigators who performed the case investigations and released case reports. This work is partly supported by the National Key Research and Development Plan (2017YFC1502901) and the National Natural Science Foundation of China (grant no. 71974017).

## References

Backer, J. A., Klinkenberg, D., & Wallinga, J. (2020). Incubation period of 2019 novel coronavirus (2019-ncov) infections among travellers from wuhan, china, 20-28 january 2020. Eurosurveillance, 25. doi:https://doi.org/10.2807/1560-7917.ES.2020.25.5.2000062.

Bui, L. V., Nguyen, T. T., & Nguyen, H. (2020). Early estimation of reproduction number of covid-19 in vietnam. medRxiv,. doi:10.1101/2020.03.28.20046136.

Du, Z., Xu, X., Wu, Y., Wang, L., Cowling, B. J., & Meyers, L. A. (2020). The serial interval of covid-19 from publicly reported confirmed cases. Emerg Infect Dis.,. doi:10.3201/eid2606.200357.

Ganyani, T., Kremer, C., Chen, D., Torneri, A., Faes, C., Wallinga, J., & Hens, N. (2020). Estimating the generation interval for covid-19 based on symptom onset data. medRxiv,. doi:10.1101/2020.03.05.20031815.

Leonhard, H., Niel, H., Philip D, O., & Jacco, W. (2019). Handbook of infectious disease data analysis. Chapman and Hall/CRC.

Li, Q., Guan, X., Wu, P., Wang, X., Zhou, L., Tong, Y., Ren, R., Leung, K. S., Lau, E. H., Wong, J. Y., Xing, X., Xiang, N., Wu, Y., Li, C., Chen, Q., Li, D., Liu, T., Zhao, J., Liu, M., Tu, W., Chen, C., Jin, L., Yang, R., Wang, Q., Zhou, S., Wang, R., Liu, H., Luo, Y., Liu, Y., Shao, G., Li, H., Tao, Z., Yang, Y., Deng, Z., Liu, B., Ma, Z., Zhang, Y., Shi, G., Lam, T. T., Wu, J. T., Gao, G. F., Cowling, B. J., Yang, B., Leung, G. M., & Feng, Z. (2020). Early transmission dynamics in wuhan, china, of novel coronavirus-infected pneumonia. New England Journal of Medicine, 0, null. doi:10.1056/NEJMoa2001316.

Linton, N. M., Kobayashi, T., Yang, Y., Hayashi, K., Akhmetzhanov, A. R., Jung, S.-m., Yuan, B., Kinoshita, R., & Nishiura, H. (2020). Incubation period and other epidemiological characteristics of 2019 novel coronavirus infections with right truncation: A statistical analysis of publicly available case data. Journal of Clinical Medicine, 9. doi:10.3390/jcm9020538.

Liu, T., Hu, J., Xiao, J., He, G., Kang, M., Rong, Z., Lin, L., Zhong, H., Huang, Q., Deng, A., Zeng, W., Tan, X., Zeng, S., Zhu, Z., Li, J., Gong, D., Wan, D., Chen, S., Guo, L., Li, Y., Sun, L., Liang, W., Song, T., He, J., & Ma, W. (2020). Time-varying transmission dynamics of novel coronavirus pneumonia in china. bioRxiv,. doi:10.1101/2020.01.25.919787.

Ma, S., Zhang, J., Zeng, M., Yun, Q., Guo, W., Zheng, Y., Zhao, S., Wang, M. H., & Yang, Z. (2020). Epidemiological parameters of coronavirus disease 2019: a pooled analysis of publicly reported individual data of 1155 cases from seven countries. medRxiv,. doi:10.1101/2020.03.21.20040329.

Moran, K. (2020). Epidemiologic characteristics of early cases with 2019 novel coronavirus (2019-ncov) disease in korea. Epidemiol Health, 42, e2020007–0. doi:10.4178/epih.e2020007.

Nishiura, H., Linton, N. M., & Akhmetzhanov, A. R. (2020). Serial interval of novel coronavirus (2019-ncov) infections. medRxiv,. doi:10.1101/2020.02.03.20019497.

Ping, K. (2020). Epidemiologic characteristics of covid-19 in guizhou, china.medRxiv,. doi:10.1101/2020.03.01.20028944.

Qian, G., Yang, N., Ma, A. H. Y., Wang, L., Li, G., Chen, X., & Chen, X. (2020).A COVID-19 Transmission within a family cluster by presymptomatic infectors in China. Clinical Infectious Diseases,. doi:10.1093/cid/ciaa316. Ciaa316.

Reich, N. G., Lessler, J., Cummings, D. A. T., & Brookmeyer, R. (2009). Estimating incubation period distributions with coarse data. Statistics in Medicine, 28, 2769–2784. doi:10.1002/sim.3659.

Song, Y., Wu, J., Thompson, R., & Shen, Z. (2020). Further improved epiestim by distinguishing imported, infected by imported, infected by local cases and its application to covid-19 in china. manuscript in preparation,.

Thompson, R., Stockwin, J., van Gaalen, R., Polonsky, J., Kamvar, Z., Demarsh, P., Dahlqwist, E., Li, S., Miguel, E., Jombart, T., Lessler, J., Cauchemez, S., & Cori, A. (2019). Improved inference of time-varying reproduction numbers during infectious disease outbreaks. Epidemics, 29, 100356. doi:https://doi.org/10.1016/j.epidem.2019.100356.

Tindale, L., Coombe, M., Stockdale, J. E., Garlock, E., Lau, W. Y. V., Saraswat, M., Lee, Y.-H. B., Zhang, L., Chen, D., Wallinga, J., & Colijn, C. (2020). Transmission interval estimates suggest pre-symptomatic spread of covid-19. medRxiv,. doi:10.1101/2020.03.03.20029983.

Wallinga, J., & Lipsitch, M. (2006). How generation intervals shape the relationship between growth rates and reproductive numbers. Proceedings of the Royal Society B: Biological Sciences, 274, 599–604.

Wang, Y., & Teunis, P. F. (2020). Strongly heterogeneous transmission of covid-19 in mainland china: local and regional variation. medRxiv,. doi:10.1101/2020.03.10.20033852.

Wei, W., Li, Z., Chiew, C., Yong, S., Toh, M., & Lee, V. (2020). Presymptomatic Transmission of SARS-CoV-2 — Singapore, January 23–March 16, 2020. MMWR Morb Mortal Wkly Rep.,. doi:10.15585/mmwr.mm6914e1.

WHO (2020). Coronavirus disease (covid-19) outbreak situation. https://www.who.int/emergencies/diseases/novel-coronavirus-2019. Accessed: 2020-4-11.

You, C., Deng, Y., Hu, W., Sun, J., Lin, Q., Zhou, F., Pang, C. H., Zhang, Y., Chen, Z., & Zhou, X.-H. (2020). Estimation of the time-varying reproduction number of covid-19 outbreak in china. medRxiv,. doi:10.1101/2020.02.08.20021253.

Zhang, J., Litvinova, M., Wang, W., Wang, Y., Deng, X., Chen, X., Li, M., Zheng, W., Yi, L., Chen, X., Wu, Q., Liang, Y., Wang, X., Yang, J., Sun, K.,Longini, I. M., Halloran, M. E., Wu, P., Cowling, B. J., Merler, S., Viboud, C., Vespignani, A., Ajelli, M., & Yu, H. (2020). Evolving epidemiology of novel coronavirus diseases 2019 and possible interruption of local transmission outside hubei province in china: a descriptive and modeling study. medRxiv,. doi:10.1101/2020.02.21.20026328.

Zhao, S., Gao, D., Zhuang, Z., Chong, M., Cai, Y., Ran, J., Cao, P., Wang, K., Lou, Y., Wang, W., Yang, L., He, D., & Wang, M. (2020). Estimating the serial interval of the novel coronavirus disease (covid-19): A statistical analysis using the public data in hong kong from january 16 to february 15, 2020. medRxiv,. doi:10.1101/2020.02.21.20026559.

